# Application of Omni-ATAC to Profile Chromatin Accessibility Before and After Ovarian Tissue Cryopreservation

**DOI:** 10.1101/2021.04.29.21256316

**Authors:** Jennifer A. Shannon, Aishwarya Sundaresan, Orhan Bukulmez, Zexu Jiao, Sarah Capelouto, Bruce Carr, Laura A. Banaszynski

## Abstract

Ovarian tissue cryopreservation and subsequent autologous transplantation has allowed resumption of endocrine function as well as fertility in certain populations. However, graft function is short-lived due to ischemia and aberrant follicular activation post-transplantation. While many studies have focused on gene expression, we wanted to determine whether cryopreservation itself had a deleterious effect on regulatory elements that might influence transcriptional integrity and graft performance. In this study, we used Omni-ATAC to assess genome-wide chromatin accessibility in primary human follicles before and after cryopreservation. Omni-ATAC from fresh ovarian follicles identified active regulatory elements expected to be functional in oocytes and granulosa cells, and gene ontology was consistent with RNA translation/processing and DNA repair. While promoter accessibility was largely maintained in cryopreserved ovarian follicles, we observed a widespread increase in the number of accessible enhancers. Transcription factor motif analysis and gene ontology suggested that this dysregulation was focused around the epithelial-mesenchymal transition. Indeed, transcription factor binding was noted in major pathways involved in this transition: TGF-β and Wnt signaling. Overall, our work provides the first genomic analysis of active regulatory elements in matched fresh and cryopreserved ovarian follicles as they undergo the process of ovarian tissue cryopreservation. Our data suggest that the process of cryopreservation activates an epithelial-mesenchymal transition state, which may lead to graft burn-out post-transplantation. Optimizing this technique in relation to this transition may therefore be an important step towards improving graft longevity and patient outcomes in fertility preservation.

**Summary sentence:** Cryopreservation of ovarian cortical tissue results in activation of differentiation and EMT pathways in follicles, which may explain graft burnout after autotransplantation.

## Introduction

Ovarian tissue cryopreservation with subsequent autologous transplantation is a promising technique that has allowed resumption of endocrine function as well as fertility in certain populations. Most successfully, the technique has been applied to patients facing gonadotoxic therapy as well as a subset of patients with premature ovarian insufficiency [1–5]. It is the only option for fertility preservation in pre-pubertal patients needing gonadotoxic therapy. Autologous transplant of cryopreserved ovarian tissue is responsible for around 200 births worldwide [6], with the first birth from cryopreserved pre- pubertal tissue occurring in 2015 [7]. Outcomes are promising, with endocrine function lasting on average 5 years [4,5,8,9], and live birth rates of up to 41% [5].

The technique of ovarian tissue cryopreservation with subsequent autologous transplantation is multi- faceted, and continued efforts focus on optimization. These efforts aim to address the best method to process the tissue, to preserve graft longevity, and to maximize graft fertility potential. Much research has occurred on a clinical scale, for example, method of cryopreservation, method of transplantation, and graft management post-transplantation [10–13]. Other research to optimize this technique has focused on the molecular mechanisms behind limited graft longevity. Longevity is dependent on functional ovarian follicles [14,15], and the majority of follicles are lost within the first few days of transplant [16,17]. Ischemia and uncontrolled follicular activation are the main causes behind this loss [16,18–20]. To address this, some have applied techniques used in skin grafting to expedite the process of neovascularization [21]. Others have focused on additive factors, like VEGF-A, vitamin E, or hyaluranon [22], or bFGF [23]. Yet another group utilized known pathways involved in follicular activation to increase follicular activation, with reported live births [2].

Our study aims to add another layer to the current understanding of the molecular pathways affected by this technique. Our goal is to apply a new method, omni-ATAC, to profile chromatin accessibility in ovarian follicles during the process of ovarian tissue cryopreservation, and to determine if cryopreservation itself has a deleterious effect on regulatory element chromatin structure and transcriptional integrity that might influence graft performance.

## Materials & Methods

### Ovarian Tissue Procurement and Processing

The Institutional Review Board of UT Southwestern Medical Center approved the use of human tissue. Written informed consent was obtained from women undergoing oophorectomy while receiving surgery for benign gynecologic indications, such as abnormal uterine bleeding or pelvic pain. Six women participated and ranged in age from 35 to 45 years old. On average, about three 1 cm ovarian slices were obtained per sample. Samples were placed in HTF media (Cooper Surgical) and transported from the operating room on ice to the laboratory for processing. Ovarian slices were sharply dissected into cortical pieces, ranging from 5-10 x 10 x 1 mm in dimension. Two pieces were submitted for histopathologic evaluation, and two pieces were used for immediate tissue digestion and follicular isolation. The remaining pieces were vitrified, and subsequently warmed, according to the previously published protocol by Silber et al. 2017 [24] with some modifications.

### Ovarian Tissue Digestion and Follicle Isolation

Digestion of ovarian cortical pieces and isolation of ovarian follicles followed previously published protocols [25,26]. For fresh isolation, the cortical pieces were sharply dissected into smaller pieces of about 2×2×2mm in size. These were added to a 50mL conical tube with 10mL Dulbecco’s phosphate-buffered saline (PBS, UTSW General Store) and 0.28 Wunsch units/mL Liberase DH (Sigma-Aldrich). The conical tube was sealed with parafilm and placed in a water bath at 37°C with gentle agitation. The digestion was terminated after 75 minutes by adding 10mL of 10% fetal bovine serum in Dulbecco’s PBS at 4°C. The supernatant was removed, and the tissue was placed on a petri dish and inspected under a Leica M165 FC stereomicroscope. Follicles were isolated from the surrounding stromal tissue via careful blunt dissection. A Cook Flexipet™ (Cook Medical) was used to pick up the follicles and transfer them into a 1.5mL Eppendorf tube with 250 uL of 10% FBS at 4°C.

For cryopreserved tissue digestion, the cortical tissue became less pliable and the follicles less prominent from the surrounding stromal tissue, so two modifications were made to aid with follicle identification. First, 150 uL of neutral red (NR, final concentration 50mcg/mL, Sigma-Aldrich) was added to the 50mL conical tube during tissue digestion [27]. Second, the McIlwain Tissue Chopper (Ted Pella) was used after digestion to further sharply dissect cortical pieces [25].

### Histology and Immunohistochemistry

The effects of cryopreservation on ovarian cortical tissue was evaluated by both histology and immunohistochemistry. Tissue was evaluated at two stages: fresh and cryopreserved. Two cortical pieces per stage were submitted for evaluation. Preparation of slides occurred through the UT Southwestern Histo Pathology Core Laboratory. The cortical pieces were fixed in formalin solution and processed into 5um slices. Images were procured at 20x magnification using a Zeiss Axioskop-2 Mot Plus microscope (Carl Zeiss, Germany) and a Leica DFC 450C camera using the Leica Application Suite v4.3.0 (Leica Microsystems, Switzerland).

Hematoxylin- and eosin-staining was used to evaluate follicle number and classification [28]. One reviewer procured images at 20x magnification. Ovarian follicles were counted and classified according to Gougeon [29] into one of three follicle stages: (i) primordial follicle; (ii) primary follicle; (iii) secondary follicle.

Immunohistochemistry was used to evaluate both proliferative and apoptotic activity in the follicles. A separate single reviewer procured fluorescent images at 20x magnification. Fluorescein (FITC - green, Thermo Fisher) was used to stain the antigen of interest, and Cy3 (bright orange, Thermo Fisher) was used as the nuclear counterstain. Images were then analyzed using the ImageJ platform (v.2.0.0-rc- 59/1.51n). The nuclear antigen Ki-67 was used to evaluate proliferative activity in the follicles [28]. Ki-67 is associated with cellular proliferation, present only during the active phase of the cell cycle (late GI, S, G2, and M phase). A proliferation index was calculated, defined as the percentage of Ki-67 positive granulosa cells over total granulosa cells for each class of follicle. Follicular apoptotic activity was evaluated using a terminal deoxynucleotidyl transferase dUTP nick end labeling (TUNEL) assay, which detects DNA fragmentation [30]. DNA fragmentation occurs in the final phase of apoptosis. An apoptotic index was calculated, defined as the percentage of TUNEL positive granulosa cells over total granulosa cells for each class of follicle.

### Omni-ATAC Transposition

Analysis of gene regulation using a modified omni-ATAC approach [31] took place on isolated follicles from two time points: fresh tissue and cryopreserved and thawed tissue. Each follicle was estimated to have about 1000 cells, and reagent concentrations were adjusted accordingly, most importantly to keep the transposase:cell number ratio similar. Following the Corces protocol, a stock resuspension buffer was made using 1M Tris-HCl pH 7.4 (final concentration 10mM), 5M NaCl (final conc. 10mM), 1M MgCl_2_ (final conc. 3mM), and molecular water.

After isolation, the follicle suspension was pelleted at 500 RCF for 5 minutes at 4°C. The supernatant was carefully removed so as to not disturb the pellet. The pellet was then resuspended in 50 uL PBS by finger tap. The suspension was then centrifuged again at 500 RCF for 5 minutes at 4°C. The supernatant was carefully removed. The lysis buffer was added (for most reactions; 48.5uL resuspension buffer, 0.5uL 10% NP-40 [final 0.1% v/v], 0.5uL 10% Tween-20 [final 0.1% v/v], 0.5uL 1% digitonin [final 0.01% v/v]), and the pellet was resuspended by finger tap. The suspension was incubated on ice for 3 minutes. A wash buffer was then added (for most reactions; 990uL resuspension buffer, 10uL 10% Tween-20 [final 0.1% v/v]), and the suspension was immediately centrifuged at 500 RCF for 10 minutes at 4°C. The supernatant was carefully removed. The transposition reaction was then added based on cell number (reaction included Illumina TDE1 Tagment DNA Enzyme, Illumina TD [Tagment DNA] Buffer, PBS, 10% Tween-20, 1% digitonin, molecular water). The pellet was suspended by finger tap, and the suspension was incubated on a thermomixer at 37°C with 1000 rpm of agitation for 30 minutes.

To stop the transposition, the suspension was subjected to immediate purification using the QIAquick PCR purification kit (Qiagen) with MinElute columns (Qiagen) [32]. The purified DNA fragments were then resuspended in 15 uL molecular water (Sigma-Aldrich), run through the column twice to ensure complete yield. To assess yield prior to library preparation, the QuBit dsDNA HS Assay (Thermo Fisher) was used to analyze the DNA concentration in 1 uL of the resulting suspension.

### Library Preparation

The PCR amplification protocol was performed as previously reported with minor adaptations [32]. The purified DNA fragments underwent an initial PCR reaction of 5 cycles (25uL NEBNext® Ultra™ II Q5® Master Mix, 7uL sample, 8uL molecular water, 5uL Nextera/Illumina Index 1 [i7] adapter, 5uL Nextera/Illumina Index 2 [i5] adapter). The resulting product was diluted 1 uL in 1000 uL molecular water and underwent quantitative PCR with Power SYBR Green, following the KAPA protocol. 6uL master mix + primer solution was added to 4uL standard or dilute sample; samples were run in duplicate using a LightCycler® 480 Instrument II system (Roche). The quantitative 2^nd^ derivative calculation was then entered into an Excel program, and the number of additional PCR cycles were calculated to achieve a final concentration of between 10-20 nM.

### Library Purification

After quantitative PCR, the libraries were purified using a double-sided bead purification protocol. The AMPure XP beads (Beckman Coulter) were warmed to room temperature and vortexed into suspension. The amplified library was transferred to a new 1.5mL Eppendorf tube, and 0.5x volume beads were added and mixed thoroughly with pipette action. The mixture was incubated at room temperature for 10 minutes, and then placed on a magnetic rack for 5 minutes. The supernatant was removed and transferred to a new 1.5mL Eppendorf tube. 1.3x volume AMPure beads were then added and mixed thoroughly with pipette action. The mixture was incubated at room temperature for 10 minutes, and then placed on a magnetic rack for 5 minutes. The supernatant was carefully removed and discarded.

The bead pellet was then washed with 200uL freshly made 80% EtOH: the volume was pipetted down the opposite side of the Eppendorf tube as the pellet, covered the pellet for 30 seconds, and was then removed. This process was repeated for a total of four washings. After the washings were completed, the Eppendorf tubes were left open for 10 minutes to allow evaporation of all the EtOH. The beads were then resuspended in 20uL molecular water. The Eppendorf tube was placed back on the magnetic rack for 3 minutes, and the supernatant containing the purified library was transferred to a new 1.5mL Eppendorf tube. The quality of the purified library was assessed using Bioanalyzer High Sensitivity Analysis on a 2200 TapeStation (Agilent).

### Sequencing

The purified library was quantified using a Qubit dsDNA HS Assay Kit (Thermo Fisher). Libraries with unique adapter barcodes were pooled, multiplexed, and sequenced on an Illumina NextSeq 500 (75 bp paired-end reads). Typical sequencing depth was at least 50 million reads per sample.

### Omni-ATAC Quality Control, Alignment, and Normalization

Quality of the ATAC-seq datasets was assessed using the FastQC tool. The ATAC-seq reads were then aligned to the human reference genome (hg19) using BWA (v.0.7.5). For unique alignments, duplicate reads were filtered out. The resulting uniquely mapped reads were normalized to the same read depth across all samples and converted into bigWig files using BEDTools [33] for visualization in Integrative Genomics Viewer [34]. The fragment length distribution plots were generated using CollectInsertSizeMetrics tool from Picard (v 2.10.3).

### Denoising

AtacWorks(v 0.3.0) was used to denoise the ATAC-seq data. All the fresh and cryopreserved and thawed samples were first preprocessed using the pipeline available at https://github.com/zchiang/atacworks_analysis/tree/master/preprocessing. After preprocessing, the atac-seq data was denoised to improve the signal-to-noise ratio using the model available from https://ngc.nvidia.com/catalog/models/nvidia:atac_bulk_lowqual_20m_20m. The denoised tracks as well as denoised peaks were obtained from atacworks. The denoised BigWig files were used to generate average ATAC-seq profiles using plotProfile tool from deepTools (v.3.4.3).

### Peak Analysis

The denoised peaks across all samples in each group were merged to create a consensus peakset using HOMER mergePeaks (v.4.9) and venn diagrams were generated using custom R scripts. Peaks were annotated to nearest genes using HOMER annotatePeaks.pl (v.4.9) with default settings.

### Classification of Regulatory Elements

*Cis*-Regulatory elements (cis-REs) like promoters, enhancers, and insulators were predicted using CoRE-ATAC.

### Motif Enrichment Analysis

Motif matching and enrichment of the open chromatin regions was performed using the motif analysis tools from Regulatory Genomics Toolbox (RGT)(v0.12.3). Motif matching of the denoised and merged peaks of fresh and thawed samples.

## Data Availability

Data files will be deposited in the Gene Expression Omnibus database under accession number GSExxx.

## Results

### Histological assessment of follicles in fresh and cryopreserved ovarian cortical tissue

Our goal was to observe the histologic effects on both fresh and cryopreserved preantral follicles as they were subjected to ovarian tissue cryopreservation. Cortical tissue was obtained, transported, and processed into cortical pieces (**Figure 1**). With regard to H&E evaluation, the follicles in fresh samples were more evenly dispersed between primordial, primary, and secondary (**Table 1**). In cryopreserved samples, the follicles had a distinct shift toward primordial classification (**Table 1**). Surprisingly, numerically more follicles were observed in the cryopreserved tissue. While large fluctuations in follicular density between patients has been observed [12], the difference observed here occurred within the primordial follicle population of samples provided by the same patient. The average age of the study participants was forty-one years old, but the samples that showed expansion of the primordial population were in those patients less than forty years old. Interestingly, an expansion in the primordial pool after cryopreservation has been previously reported [10].

**Table 1.** Gougeon Classification of H&E-stained Fresh and Thawed Ovarian Critical Tissue

| **Table 1: Gougeon Classification of H&E-stained Fresh and Thawed Ovarian Cortical Tissue** |  |  |  |  |  |
| --- | --- | --- | --- | --- | --- |
| **Tissue Type** | **Specimens, n** | **Follicles, n** | **Proportion of follicles n (% total number)** |  |  |
|  |  |  | **Primordial** | **Primary** | **Secondary** |
| Fresh | 6 | 19 | 6 (31.6%) | 4 (21%) | 9 (47.4%) |
| Thawed | 5 | 94 | 68 (72.3%) | 10 (10.6%) | 16 (17%) |

**Figure 1.**
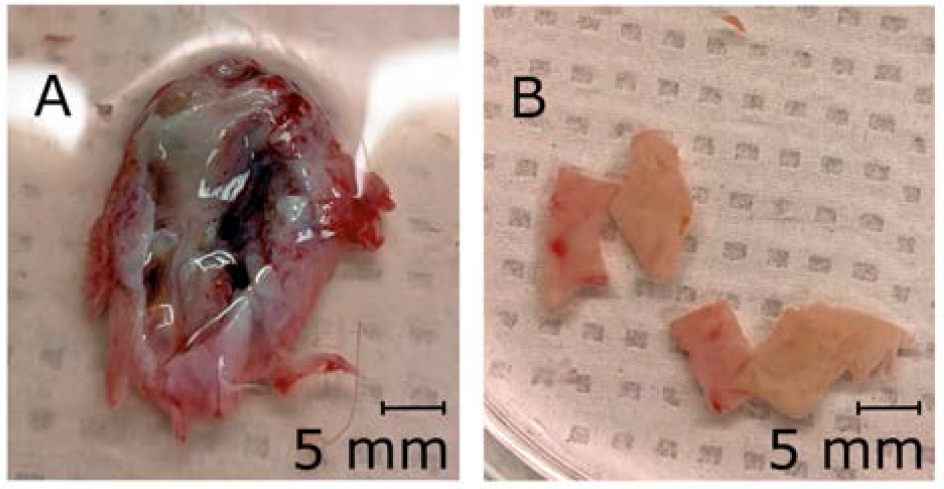
Fresh ovary processing. A) Tissue was obtained as ∼8 mm diameter ovarian slices. B) Ovarian cortical pieces, measuring 5-10 x 10 x 1 mm in dimensions, were created.

To determine the effects of cryopreservation on tissue integrity in our hands, we assessed both proliferation and apoptosis from fresh and cryopreserved tissue. No difference in proliferation was seen among fresh and cryopreserved ovarian follicles classified as either primordial or primary (**Figure 2A**). Secondary ovarian follicles did show a significant trend toward increased proliferation in the fresh state (**Figure 2A**). With regard to apoptosis, primordial and primary follicles demonstrated no difference in staining, however secondary follicles did show increased apoptosis after cryopreservation and thaw (**Figure 2B**). These data are consistent with prior evidence suggesting that primordial follicles more easily survive the cryopreservation process [35].

**Figure 2.**
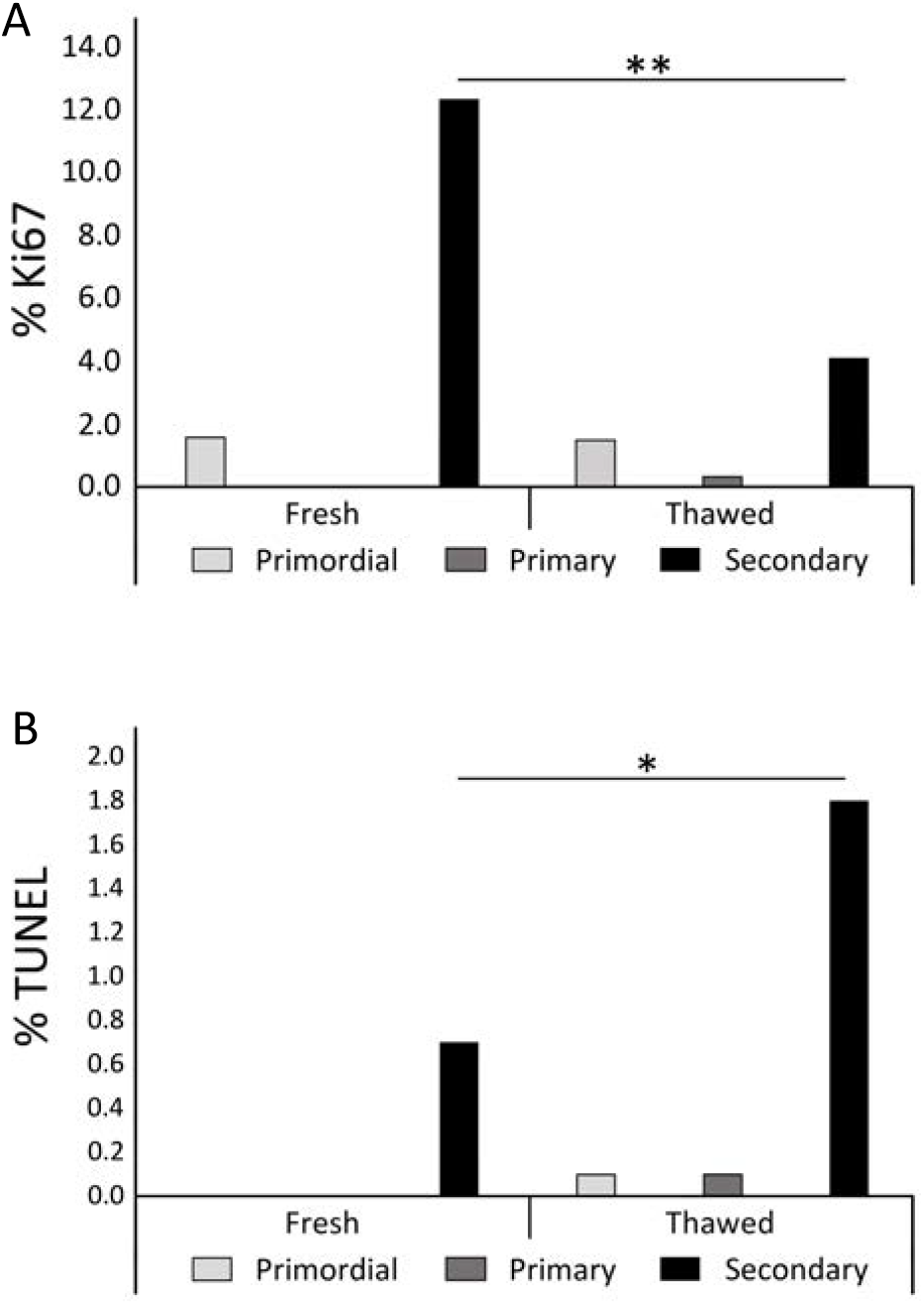
Evaluation of proliferative and apoptotic markers before and after cryopreservation. A) Percent of Ki67-positive granulosa cells out of total granulosa cells. B) Percent of TUNEL-positive granulosa cells out of total granulosa cells. Significant differences were seen between secondary follicles in the fresh and cryopreserved states (*p < 0.05, ** p < 0.001).

The addition of neutral red allowed for easier visualization (**Figure 3A**) and for the distinction between viable and non-viable follicles (**Figure 3A** versus **3B**) [27]. The improved visualization also allowed for a significant increase in follicular yield (**Figure 3C**). Overall, our immunohistochemistry data demonstrate that we isolated viable tissue. Further, preantral follicles seemed relatively unaffected histologically by cryopreservation, suggesting that they might be a good target for the genomic analyses described below.

**Figure 3.**
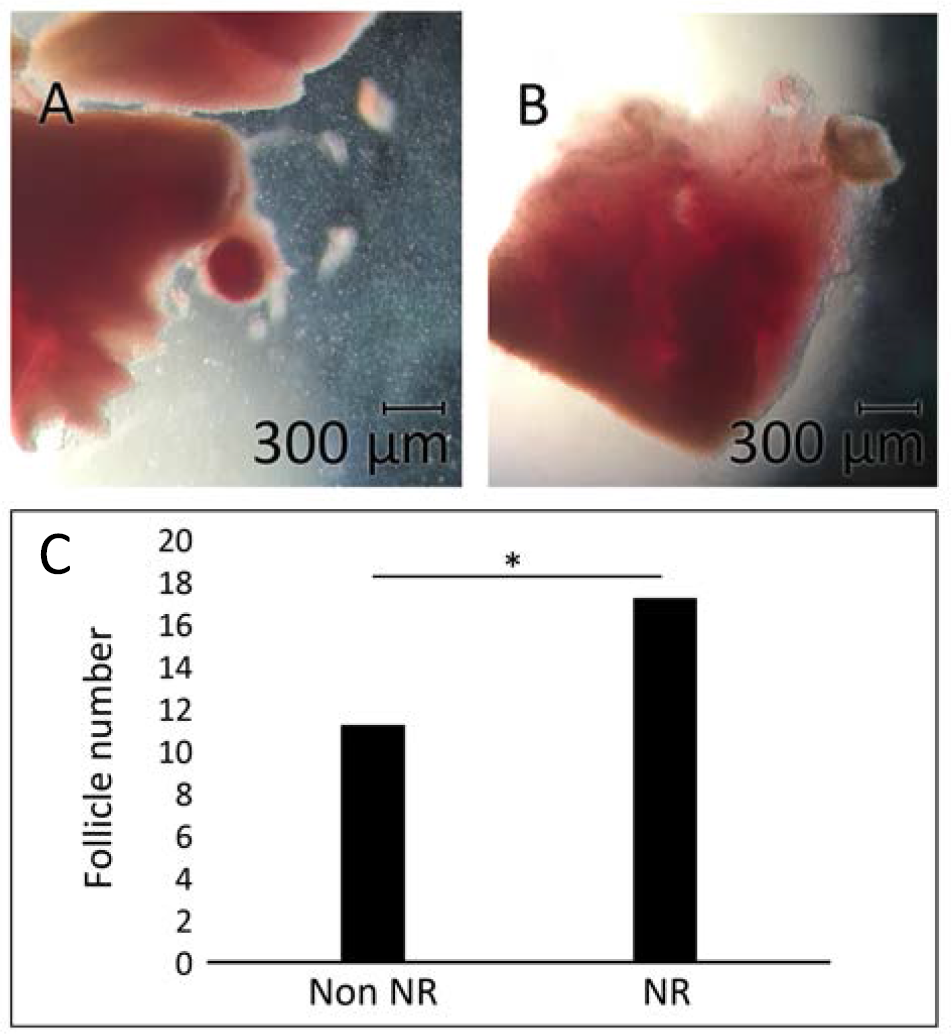
Neutral red versus non-neutral red follicle isolation. A) Neutral red-stained secondary follicle. B) Non-staining secondary follicle after incubation. C) Average follicle count per isolation (p < 0.05).

### ATAC Analysis From Fresh Ovarian Follicles

Before we could determine the effects of cryopreservation on the chromatin state at regulatory elements, we first needed to define these regions from fresh cortical tissue. Preantral follicles were isolated and subjected to Omni-ATAC, an ATAC-seq protocol that allows chromatin accessibility profiling from frozen tissue samples [31]. The insert size distribution of sequenced fragments had clear nucleosomal periodicity of around 200 bp, as expected (**Figure 4A**). Importantly, data sets obtained from individual donor samples processed immediately after dissection were highly correlated (**Figure 4B**). We observed enrichment of open chromatin at known oocyte-specific and granulosa cell-specific markers, GDF-9 and FOXL-2, respectively, that appeared consistent between donors (**Figure 4C**), indicative of isolation of the cell types of interest. To improve the signal over background in our data sets, we used a deep learning toolkit called AtacWorks [36] to denoise our sequencing data (**Figure 4D**).

**Figure 4.**
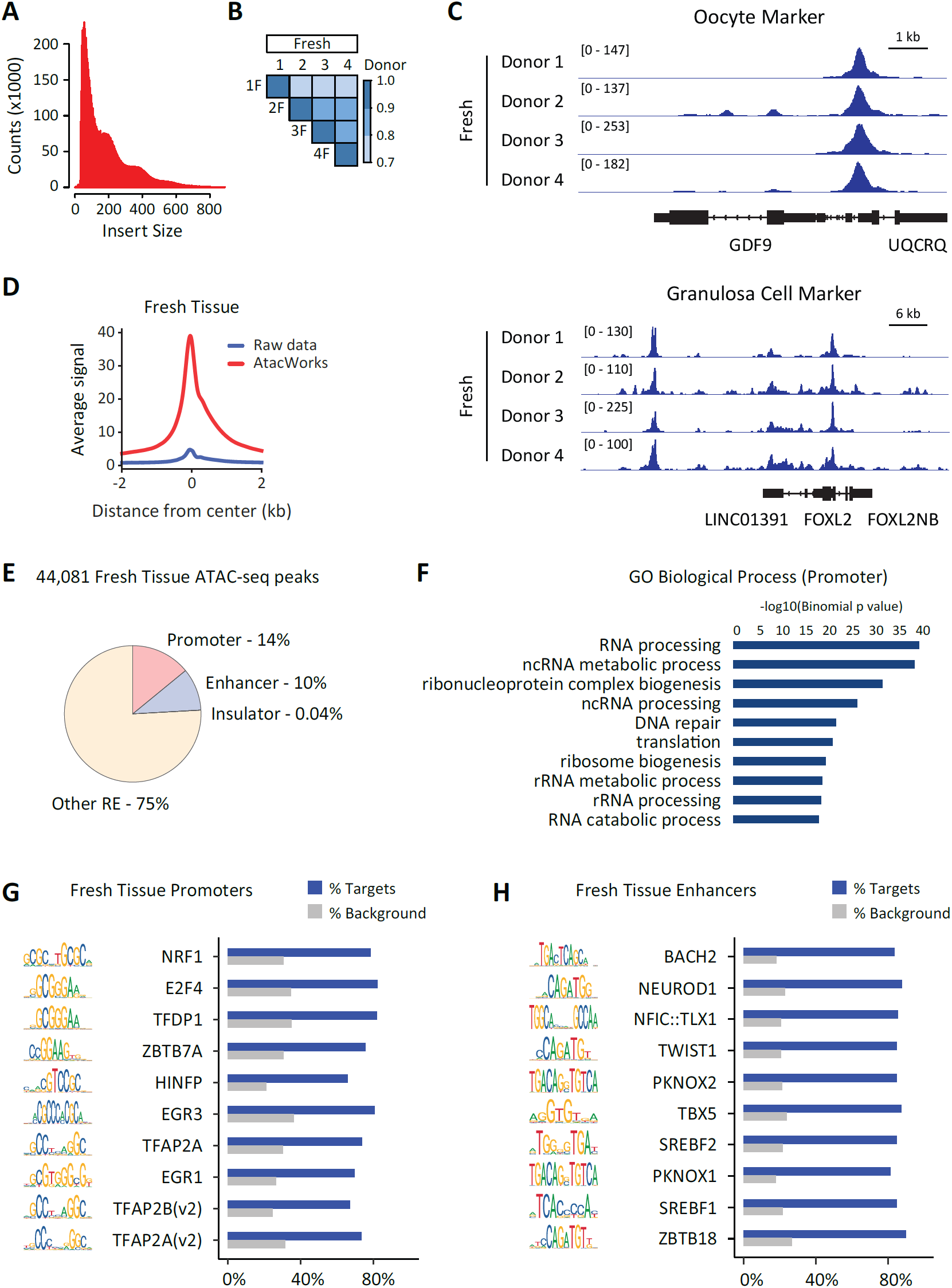
Identification of active regulatory elements in human follicles. A) ATAC-seq fragment sizes generated from human follicles. B) Correlation between ATAC-seq datasets generated from human follicles from multiple donors. C) Genome browser representations of ATAC-seq data from human follicles. Representative oocyte marker (top panel, GDF9) and granulosa cell marker (bottom panel, FOXL2) are displayed. The y axis represents read density in reads per kilobase per million mapped (RPKM) reads. D) ATAC-seq average profiles from human follicles representing enriched regions before (Raw data) and after (AtacWorks) denoising using AtacWorks. E) Functional annotation of ATAC-seq peaks as promoter, enhancer, insulator, or other regulatory element (RE) using CoRE-ATAC. F) Gene ontology analysis of biological processes associated with active promoters identified from follicle ATAC- seq data. G and H) Motif enrichment for (G) follicle promoters and (H) follicle enhancers identified from ATAC-seq data. Top 10 motifs based on enrichment over background are displayed.

We next determined regions of accessible chromatin from our fresh samples and used a deep learning framework called CoRE-ATAC to functionally assign the identified regulatory elements as promoters, enhancers, or insulators [37]. Using this method, 14% of the identified regulatory elements were annotated as promoters and 10% were annotated as enhancers (**Figure 4E**). We then used gene ontology to identify biological processes associated with the active promoters we identified from fresh follicles (**Figure 4F**). Overall these regulatory regions were associated with the translation and processing of RNA, as well as some DNA repair. These observations are in line with the reliance of the oocyte on translation as a mechanism of gene regulation rather than transcription [38]. To understand the regulatory networks active in preantral follicles, we used HINT-ATAC to identify the top 10 motifs enriched in the ATAC-seq data over background for both promoters and enhancers. At promoters, we identify motifs for transcription factors reported to play a role in germ cell biology, e.g., NRF1 and TFAP2A [39–41] (**Figure 4G**). We also observe a number of motifs for transcription factors associated with cell cycle regulation (e.g., E2F4, TFDP1, HINFP), thought to play an important role in establishing cell cycle post-fertilization and in early embryonic cells [42,43]. Interestingly, at enhancers, we identified motifs for a number of homeobox transcription factors (e.g., TLX1 and PKNOX1/2). While homeobox proteins such as the HOX genes are typically associated with developmental patterning, there are reports of oocyte-specific homeobox proteins [44], in support for a role for this class of proteins in the oocyte. We also identify the TWIST1 motif in preantral follicle enhancers, in line with the epithelial-to-mesenchymal (EMT) transition having a role during folliculogenesis [45]. In addition, we also observed motifs associated with early embryonic differentiation, such as NEUROD1 and TBX5 (**Figure 4H**). These data suggest that the oocyte and supporting granulosa cells may carry a “poised” enhancer signature [46,47] to promote lineage-specifying transcription programs upon fertilization and subsequent differentiation.

### ATAC Analysis From Cryopreserved Ovarian Follicles

Studies using cryopreserved tissue in autologous transplant demonstrate that these grafts experience follicular “burn-out” over time, in part attributed to ischemia and aberrant follicular activation post-transplant [16,48,49]. However, it remains formally possible that the act of cryopreservation itself may alter gene regulatory networks in follicles, leading to follicular exhaustion upon transplant. To determine the effect of cryopreservation on gene regulatory networks, we subjected preantral follicles from cryopreserved tissue to the same Omni-ATAC protocol described above. We obtained data from three patient samples, with two of these samples directly matched from our fresh data sets (from donors 1 and 4). Overall, data sets from cryopreserved follicles were highly correlated among themselves and also with datasets obtained from freshly dissected follicles (**Figure 5A**). ATAC-seq data sets from cryopreserved tissue were slightly lower quality compared to data sets obtained from fresh tissue, a known issue when processing frozen samples and tissues specifically [31]. Processing with the AtacWorks algorithm [36] resulted in improved signal quality, similar to what we observed with processed data from fresh tissue (**Figure 5B**).

**Figure 5.**
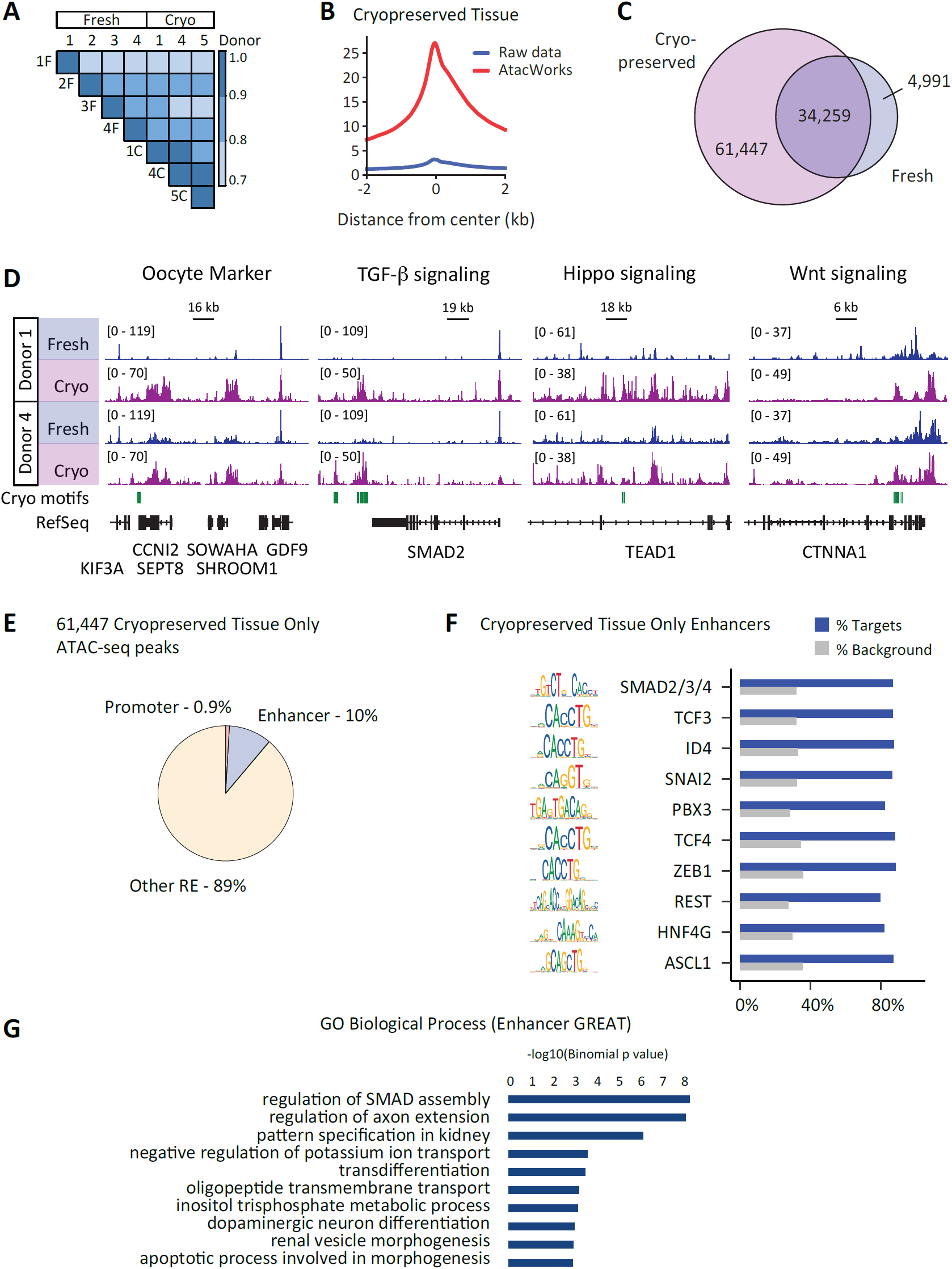
Comparison of active regulatory elements in follicles before and after cryopreservation. A) Correlation between ATAC-seq datasets generated from human follicles from multiple donors before (Fresh) or after (Cryo) cryopreservation. B) ATAC-seq average profiles from human follicles subjected to cryopreservation representing enriched regions before (Raw data) and after (AtacWorks) denoising using AtacWorks. C) Venn diagram representing overlap between ATAC-seq enriched regions identified from fresh and cryopreserved human follicles. D) Genome browser representations of ATAC-seq data from human follicles before and after cryopreservation. The y axis represents read density in reads per kilobase per million mapped (RPKM) reads. E) Functional annotation of ATAC-seq peaks identified only from cryopreserved follicles using CoRE-ATAC. F) Motif enrichment for follicle enhancers identified only after cryopreservation. Top 10 unique motifs present only after cryopreservation from the top 30 motifs identified from cryopreserved follicle enhancers. G) Ontology analysis of associated biological processes for putative target genes (using GREAT) of enhancers identified from follicles only after cryopreservation.

We next wanted to compare the open chromatin landscape between fresh and cryopreserved preantral follicles. First, we find that the majority of peaks identified from fresh follicles are still present in cryopreserved follicles (34,259/39,250 or 87%, **Figure 5C**), demonstrating that cell identity is maintained through cryopreservation. However, we observe a dramatic increase in the number of open chromatin regions present in follicles after cryopreservation (95,706 peaks from cryopreserved follicles vs. 44,081 peaks from fresh follicles, **Figure 5C**). This increased accessibility was evident in direct comparison of specific loci from data sets obtained from donor-matched fresh and cryopreserved samples (**Figure 5D**). CoRE-ATAC was then used to functionally assign open chromatin regions identified only after cryopreservation (n=61,447). Surprisingly, whereas 10% are annotated as enhancers, only 1% of these regions are annotated as promoters (**Figure 5E**). This is in contrast to results from fresh follicles, where around 14% of the regulatory elements identified are annotated as promoters. This data suggests that, in general, while the open chromatin state at promoters is unaffected by cryopreservation, this process results in dysregulation of chromatin states leading to increased DNA accessibility specifically at enhancers.

We then considered two possible mechanisms by which enhancers could be dysregulated by cryopreservation. First, this could represent a stochastic process, by which chromatin structure is damaged by the process of cryopreservation and the affected enhancer elements are random. Second, we imagined that the act of cryopreservation might affect specific pathways, which would be evidenced by the presence of unique motifs when considering enhancers identified only from the cryopreserved follicles. To determine whether unique transcription factor motifs were present in cryopreserved follicles, we considered the top 30 enriched motifs in either all fresh enhancers or only cryopreserved enhancers. We identified 19 motifs that were common to both sets of enhancers, and 11 motifs that were unique to each group. Of these, we present the top 10 motifs that were enriched in enhancers identified only in cryopreserved follicles (**Figure 5F**). Included are motifs associated with well-known ovarian signaling pathways, for example the SMAD family involved in TGF-β signaling and the TCF family involved in Wnt signaling. Both the TGF-β and Wnt signaling pathways are involved in follicular activation [50,51]. Further, we identified motifs that are targets of these signaling pathways that have known roles in EMT (e.g., ID4, SNAI2, ZEB1), which has been implicated in both normal physiologic and abnormal pathophysiologic processes in the female reproductive tract [45]. Finally, we performed ontology analysis of nearest neighboring genes to dysregulated enhancers using GREAT [52] (**Figure 5G**). These terms suggest that cryopreserved follicles are activating enhancers associated with TGF-β signaling (e.g., SMAD activity) and cellular differentiation. Overall, motif assessment and ontology analysis suggests that cryopreserved tissues may be responding to specific signaling pathways, and that the enhancer dysregulation observed may be specific.

## Discussion

Overall, our work provides the first genomic analysis of active regulatory elements in matched fresh and cryopreserved ovarian preantral follicles as they undergo the process of ovarian tissue cryopreservation. We find that, while chromatin accessibility appears unperturbed at promoters, cryopreserved follicles show evidence of chromatin dysregulation of enhancer regions. Signaling pathways associated with follicular activation and the epithelial-mesenchymal transition (e.g. TGB-β and Wnt signaling) appear to be activated in cryopreserved tissue, suggesting that the process of cryopreservation itself may contribute to graft burn-out and fibrosis post-transplantation.

Independent of our analysis of the effects of cryopreservation, our study is of note because it is the first to identify active regulatory elements from human follicles. Analysis of the top 10 enriched promoter motifs identified potential roles for a number of transcription factors that have previously been associated with oocyte function. For example, TFDP1, E2F4, and HINFP are all associated with cell cycle and have been reported to be maternally stored in the oocyte to facilitate rapid cell cycle activation upon fertilization [42,53]. NRF1 is associated with mitochondrial respiration and has also been implicated in oocyte activation post-fertilization [39]. Further, NRF1 may function to regulate genetic stability through regulation of the kinetochore and spindle assembly [54], dysfunction of which can occur in aged oocytes [55]. Interestingly, EGR1 has been reported as a marker of ovarian aging in mouse models [56], which may be reflective of the relatively advanced age of our donor group at an average of forty-one years old. When we consider transcription-factor binding motifs enriched at enhancers identified in follicles, we observe a number of homeobox proteins, including TLX1 and PKNOX1/2. Several of these factors have previously been observed in mouse follicles [44,57], although their function in oocyte maturation and granulosa cell differentiation remains poorly understood. Additionally, we observe motifs such as TBX5 and NEUROD1, transcription factors associated with differentiation. These data suggest that the oocyte may be primed for development, as has previously been observed in both mouse and human embryonic stem cells [46,47].

With regard to our cryopreserved data, we observed evidence of signaling through well-known ovarian signaling pathways, specifically the TGF-β, Wnt, and Hippo signaling pathways. Interestingly, these pathways are not only involved in follicular activation [58–60] but also in the epithelial-mesenchymal transition. Perhaps it is not surprising that mechanisms of cellular plasticity appear to be activated in tissue undergoing extreme mechanical and metabolic stress as a result of dissection and cryopreservation. EMT encompasses three types of cellular transitions: type 1 EMT occur predominantly during embryo- and organo-genesis, type 2 occur during episodes of tissue healing and fibrosis, and type 3 occur during carcinogenesis [61,62]. The transitory nature of EMT has led some to consider that the plasticity of this state may be due to epigenetic master regulators with widespread effects on gene expression [63]. Indeed, two of these master regulators were identified upon analysis of the top enhancer motifs: ZEB1 and SNAI2 (SLUG) [62]. These regulators are acted upon by a variety of signaling cascades, including TGF-β and Wnt.

In relation to EMT, TGF-β signaling causes repression of the epithelial E-cadherin in favor of the expression of the mesenchymal N-cadherin [62,64], a major step in the initiation of the epithelial-mesenchymal transition. There is evidence that Wnt signaling can play a role in regulating EMT, as beta-catenin has been shown to directly bind to the promoters of ZEB1 and SNAI2 (SLUG) to induce their expression [65]. Interestingly, we observe increased regulatory element engagement on TEAD1, a critical transcription factor in the Hippo signaling pathway. Hippo inhibition has been shown to reduce TGF-β signaling activity [66], suggesting that Hippo pathway may be upstream of TGF-β signaling during response to cryopreservation. High levels of TGF-β have been associated with fibrosis in the ovary [45], and graft fibrosis is one of the reasons ovarian tissue cryopreservation and auto-transplantation is not a preferred method for older women [67]. Indeed, when we attempted to xenotransplant human ovarian tissue sections into immunocompromised mouse models, we observed long-term fibrosis four weeks after transplant (data not shown). Overall, these observations suggest a complicated molecular cascade in which follicles may be responding to the mechanical and metabolic stress of cryopreservation through dysregulation of TGF-β and Wnt signaling.

Our data has a few limitations that merit consideration. First, due to the limited amount of tissue available for follicle isolation, follicles were obtained as a collective group, rather than in stage-specific groups. Although most follicles isolated were anticipated to be from a primary to early secondary stage due to follicle size and cortical location, cryopreserved samples yielded the occasional primordial nest. Thus, it is formally possible that the expansion of enhancer signals observed from cryopreserved tissue could be caused by the isolation of proportionally more primordial follicles in the cryopreserved cortex as compared to the fresh cortex. Second, again due to collection limitations, our tissue was collected from donors of more advanced age compared with many previous studies. On a broad scale, given the trauma to the tissue, it is generally plausible that the epithelial-mesenchymal transition is initiated.

However, given that many fertility-related outcomes depend on the age of the woman, one would suspect that the balance of the signals in EMT networks would also be affected by age. Perhaps younger tissue is able to survive EMT activation with less fibrosis, leading to improved graft viability. The question then becomes, Is there a way to pharmacologically tip the balance away from EMT and fibrosis by suppressing TGF-β? This strategy has been looked at with favorable results in prior mammalian studies involving end stage renal disease, renal allotransplant, peritoneal fibrosis, and pulmonary fibrosis [68–71]. Third, our cryopreservation protocol used vitrification. There have been conflicting studies regarding vitrification of ovarian tissue, with most of the live births originating from slow-freeze tissue [4,10,72–74]. Whether early differences between slow freezing and vitrification protocols affect final graft outcome is still a question under active investigation, and the use of vitrification for our protocol may not apply to tissue that has undergone slow freeze protocols.

Overall, our research provides a first glimpse into the chromatin accessibility profile of gene regulatory elements in ovarian follicles as they undergo the process of ovarian tissue cryopreservation. While promoter regions were mostly maintained through cryopreservation, enhancer regions were dysregulated in a way that suggested activation of folliculogenesis and the epithelial-mesenchymal transition. With regard to OTC, this new knowledge gives another avenue for development of interventions that could flip the transition away from follicular activation and fibrosis to preserve tissue integrity and function.

## Data Availability

Data files have been deposited in the Gene Expression Omnibus database.

## Acknowledgements

The authors thank Dr. Kaitlin Doody for her work to microscopically record the H&E ovarian cortical tissue images.

## Author Contributions

J.S. conceived the project and performed the experiments. A.S. analyzed the ATAC-seq data. Z.J. provided invaluable experience in the cryopreservation and thaw of ovarian cortical tissue. S.C. provided microscopic recording as well as analysis of immunohistochemically-stained ovarian cortical tissue. O.B., B.C., and L.B. conceived the project and provided funding for the work. J.S. and L.B. wrote the manuscript with input from all authors.

